# Sensitive detection of *Plasmodium vivax* malaria by the rotating-crystal magneto-optical method in Thailand

**DOI:** 10.1101/2021.05.13.21257180

**Authors:** Ágnes Orbán, Rhea J. Longley, Piyarat Sripoorote, Nongnuj Maneechai, Wang Nguitragool, Adam Butykai, Ivo Mueller, Jetsumon Sattabongkot, Stephan Karl, István Kézsmárki

## Abstract

The rotating-crystal magneto-optical detection (RMOD) method has been developed for the rapid and quantitative diagnosis of malaria and tested systematically on various malaria infection models. Very recently, an extended field trial in a high-transmission region of Papua New Guinea demonstrated its great potential for detecting malaria infections, in particular *Plasmodium vivax*. In the present small-scale field test, carried out in a low-transmission area of Thailand, RMOD confirmed malaria in all samples found to be infected with *Plasmodium vivax* by microscopy, our reference method. Moreover, the magneto-optical signal for this sample set was typically 1–3 orders of magnitude higher than the cut-off value of RMOD determined on uninfected samples. Based on the serial dilution of the original patient samples, we expect that the method can detect *Plasmodium vivax* malaria in blood samples with parasite densities as low as ∼ 5 − 10 parasites per microliter, a limit around the pyrogenic threshold of the infection. In addition, by investigating the correlation between the magnitude of the magneto-optical signal, the parasite density and the erythrocytic stage distribution, we estimate the relative hemozoin production rates of the ring and the trophozoite stages of *in vivo Plasmodium vivax* infections.

## Introduction

Although humanity is continuously facing novel medical challenges, the management of preventable infectious diseases, which place heavy burden on tropical countries, has been a long-sought goal of the WHO [1]. One such disease is malaria, and by virtue of increased global and local efforts significant progress has been made towards its control worldwide. Since many countries are working towards the substantial reduction of disease burden, the development of sensitive, rapid, high-throughput and low-cost malaria diagnostic methods applicable for mass-screening is a pressing issue in tropical diseases research [2].

One promising diagnostic approach is the utilisation of the magnetic properties of *Plasmodium*-infected red blood cells, as it may enable the rapid and quantitative detection of the infection at low cost [3–7]. During the intraerythrocytic cycle the parasites break down the hemoglobin of their host cell and sequester its iron content into the inert, submicron-sized paramagnetic hemozoin crystals, which are distinguishing features of all blood-stage *Plasmodium* infections [8, 9]. In the recent years, our research group has developed a new technique, the rotating-crystal magneto-optical diagnostic detection (RMOD) method, which is capable of the quantitative detection of malaria by measuring the amount of hemozoin produced during the course of the infection. The diagnostic capability of RMOD was tested in several steps using synthetic *β*-hematin crystals, *P. falciparum* cell cultures and mouse infections [10–13]. Furthermore, it proved to be an efficient tool for conducting rapid, yet sensitive drug susceptibility assays [14].

Recently, a detailed evaluation of RMOD has been carried out in an extended field trial in Papua New Guinea (PNG) involving almost one thousand malaria-suspected patients and multiple reference methods [15]. One important — and anticipated — observation of the study was that infections with different species led to different average blood hemozoin levels, resulting in a more sensitive detection of *P. vivax* than *P. falciparum*. However, the presence of stage V *P. falciparum* gametocytes increased the likelihood of identifying *P. falciparum* infections due to their high hemozoin content. As another key finding, the mean value and the standard deviation of the magneto-optical (MO) signal for samples which tested negative with all reference methods was significantly higher than expected from previous RMOD measurements on malaria-naïve blood samples [11, 12]. Accordingly, we concluded that in high-transmission settings, such as the study site in PNG, elevated hemozoin levels are likely to be maintained in the peripheral blood of a large proportion of the general population, either from concurrent low-level infections that are otherwise undetectable, or from previously resolved infections [16, 17].

In order to further investigate these observations and to assess the sensitivity of RMOD for diagnosing *P. vivax* infections in low-transmission settings, here we use the technique to detect malaria in samples confirmed to be *P. vivax*-positive by microscopy from Thailand, a country with overall low malaria endemicity [18, 19]. In addition to testing the sensitivity of the method, we evaluate the correlation between the magnitude of the MO signal and the parasite stage distributions of the samples.

The potential of the magneto-optical approach for diagnosing malaria has also been demonstrated in a recent field test by another hemozoin-based tool, Gazelle, which showed a sensitivity of 96.2% for the detection of *P*.*vivax* infections [20]. The main difference between RMOD and Gazelle is that the former detects the periodic modulation of light transmission induced by the hemozoin crystals co-rotating with a rotating magnetic field, while the latter measures light transmission in stationary states with and without a static magnetic field. In spite of this technical difference, the physical principle of detection is common. Thus, the magneto-optical quantification of hemozoin should support not only a qualitative, but a quantitative diagnosis in both cases. While this capability has not been investigated for Gazelle, it is an important objective of the present study.

## Results

### The sensitivity of RMOD in detecting *P. vivax* infections

The parasite densities of the original 35 patient samples ranged from 34 µl^−1^ to 5142 µl^−1^ with a median of 875 µl^−1^. As seen in Fig.1A., this range was well covered by the samples, though their parasite density distribution was not uniform.

**Figure 1.**
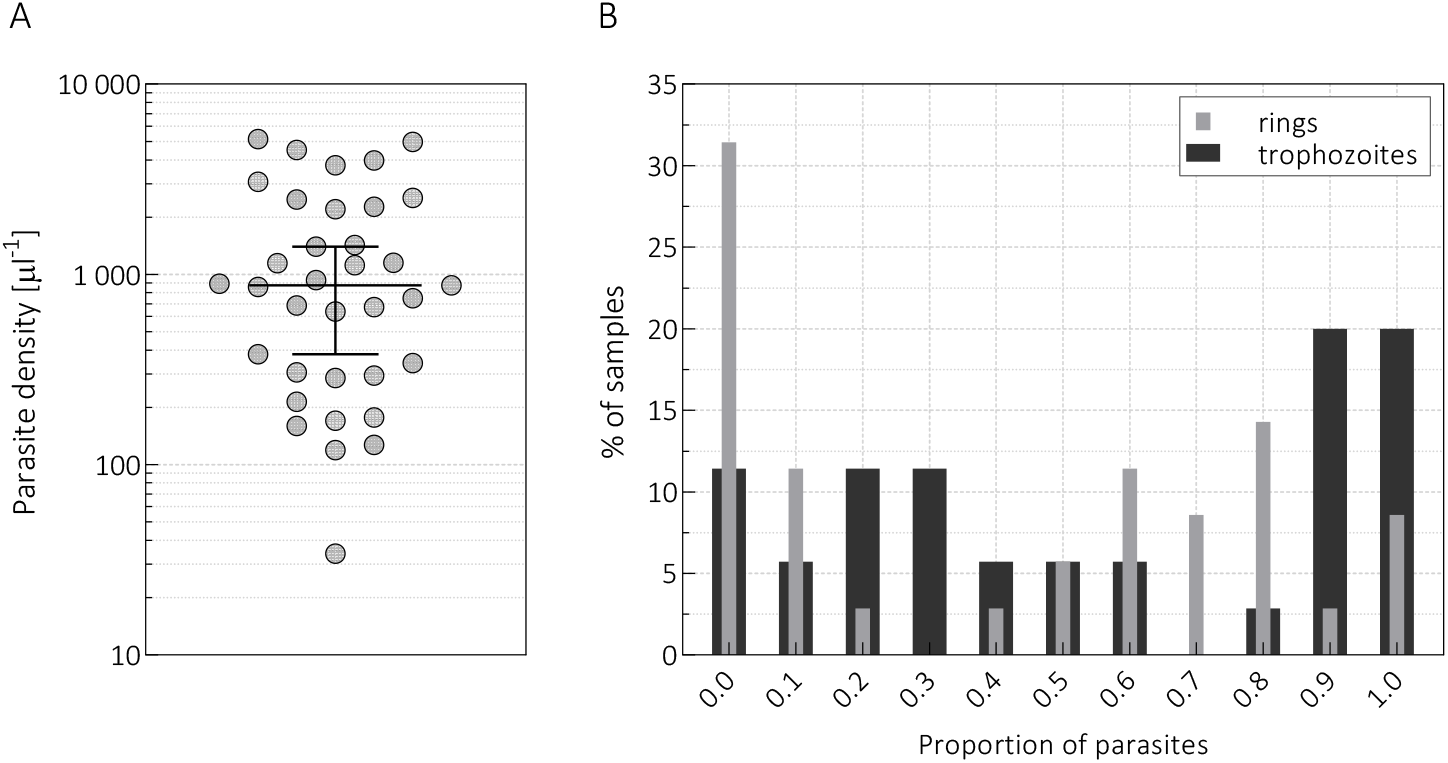
**Panel A**. The parasite density values of the *P. vivax*-infected samples (n=35) with their median and the 95% confidence interval (95%-CI) of the median. **Panel B**. The percentage of samples containing the indicated proportion of rings (light grey columns) and trophozoites (dark grey columns), i.e., histograms of the incidence rate of rings and trophozoites.

Besides the overall parasite density, the erythrocytic stage distribution was also determined by an expert microscopist, and the parasites were classified as *rings, trophozoites, schizonts, male* and *female gametocytes*. The latter three forms contributed less than 10% to the overall parasite density in all samples with one exception, in which their proportion reached 35%. The sequestration of schizonts is a well-known phenomenon in *P. falciparum* infections, but the statistics of our *P. vivax* samples suggests that it also occurs for *P. vivax* [21]. Accordingly, the blood smears exhibited mostly rings and trophozoites, but their relative ratios varied in the sample population. A large fraction of samples (n=27) showed a relatively synchronous character and contained either predominantly (≥ 70%) rings or trophozoites as seen in Fig.1B. In contrast to the dominance of ring-stage parasites in the peripheral circulation of *P. falciparum* infections, in the case of these *P. vivax*-positive samples more than 30% of the samples showed no ring forms, and 88% contained trophozoites in more than 10%.

As a first step, the level of the weak residual MO signal, that is present even in samples of malaria-naïve individuals, was measured on a pool of uninfected samples in order to establish a threshold that separates malaria-positive and negative RMOD incidences. Measurements on fourteen uninfected samples obtained from malaria-naïve individuals in Thailand, each measured in duplicates, yielded an average residual MO value of (0.50*±*0.22)mV*/*V as presented in Fig.2A. The MO signal level that separates malaria-positive and -negative incidences, i.e., the cut-off level, was determined as the mean plus two times the standard deviation of these residual values, assuming that the MO values of uninfected samples are normally distributed random variables. This led to a cut-off level of 0.94 mV/V as indicated by the horizontal black dotted line in Figs. 2B. and 2C. As displayed in Fig.2B., all the *P. vivax*-infected blood samples tested above the cut-off, yielding a 100% sensitivity on this sample set. Moreover, for more than 90% of the samples the MO values are at least one order of magnitude larger than the cut-off, implying that the method can detect lower parasite densities. To model such cases, we chose ten of the original blood samples and prepared dilution series from them using uninfected whole blood as dilution medium. The parasite densities of the original ten samples change between 34 µl^−1^ and 4980 µl^−1^, while the parasite densities of the most diluted samples cover the range of 0.0003 − 9 µl^−1^.

**Figure 2.**
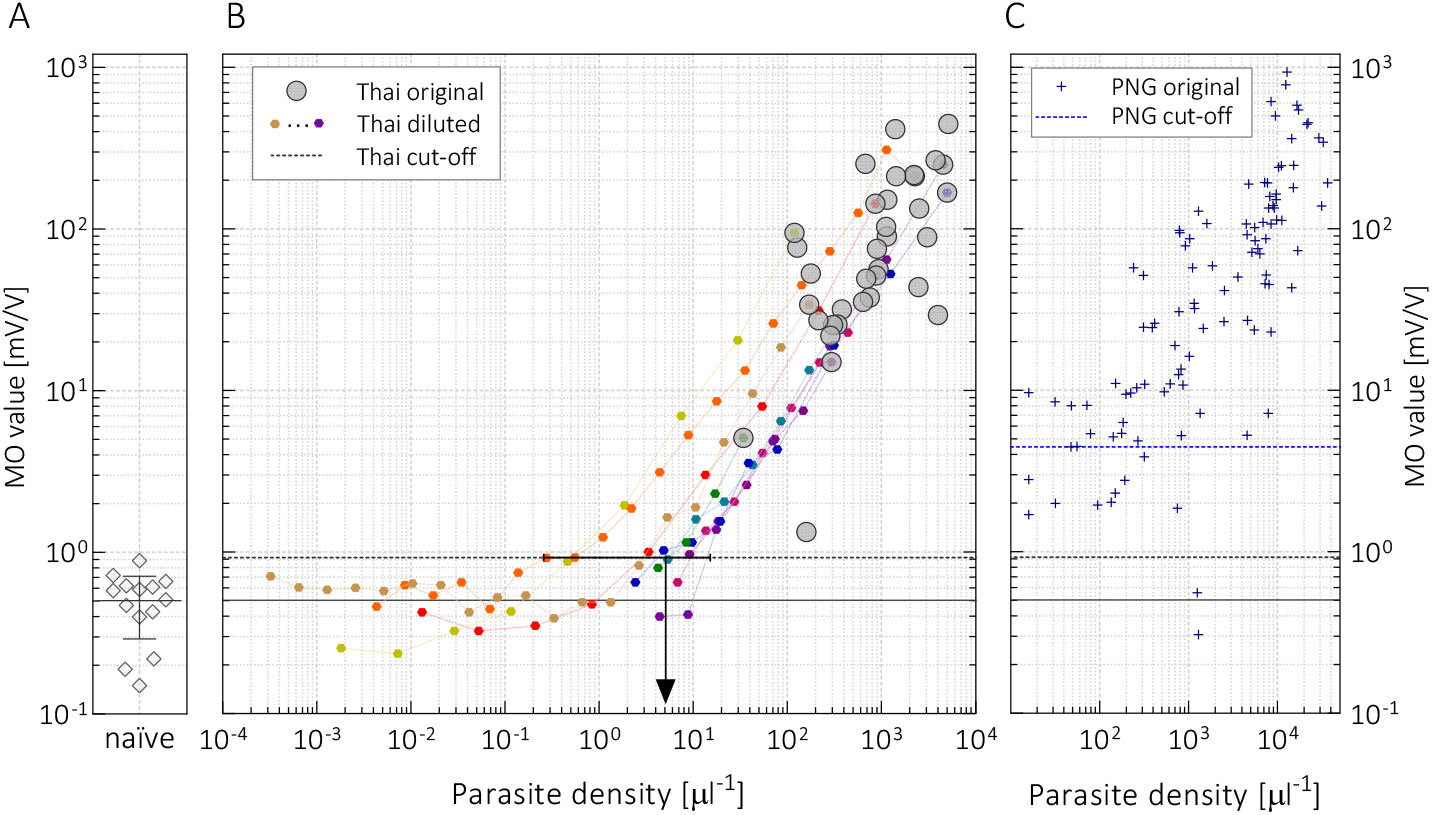
**Panel A**. The MO values of fourteen malaria-naïve samples obtained from two individuals at various time-points during the course of the study with their mean and standard deviation. **Panel B**. The MO values of the infected samples (grey circles, n=35) plotted as the function of their parasite densities. Each data point represents the average MO value of at least two technical duplicates (individual errors are below 10% and obscured by the logarithmic scale). The coloured symbols show the MO values of samples obtained by serial dilution of ten infected samples. The black continuous and dashed lines indicate the mean MO value of the malaria-naïve samples and the cut-off level, respectively, while the black arrow marks the estimated lowest detectable parasite density by RMOD. **Panel C**.The blue asterisks show the MO values of *P. vivax*-infected samples collected in Papua New Guinea (n=104), as reproduced from Ref. [15]. The blue dashed line marks the cut-off value for RMOD determined by receiver operating characteristic (ROC) analysis in that study [15].

The MO values of the diluted samples prepared from the same infected specimen (identically coloured circles in Fig.2B.) decrease linearly with increasing dilution over several orders of magnitude as their parasite densities decrease proportionally with the dilution. However, the MO values of highly diluted samples with very low parasite densities (< 1 µl^−1^) are independent of the nominal parasite load. These residual MO values are scattered around the mean residual value obtained for the uninfected controls, below the cut-off level defined previously. This further supports that this residual MO signal is unrelated to parasites. Accordingly, the limit of detection in terms of parasite density is defined as the average parasite density for samples with MO values near the cut-off, i.e., in the range of 0.94 *±* 0.08 mV*/*V. This estimate leads to a detection limit of approx. 5 µl^−1^ (range: 0.2 − 12 µl^−1^) as indicated by the black arrow (line segment) in Fig.2B. For comparison, this sensitivity threshold is somewhat better than the threshold reported in the recent field test of the Gazelle device, where the parasite density of false negative cases was reported to range from 18 to 174 µl^−1^, when optical microscopy was used as a reference [20].

### The correlation between the MO values, the parasite density and the stage distribution

As observed in Fig.2., the relationship between the parasite densities and the corresponding MO values is linear (Spearman Rank: *R* ≥ 0.95, *p <* 0.0001) for samples obtained by serial dilution as long as their MO values are above the cut-off level. However, the correlation between the MO values and parasite densities for the 1original (undiluted) samples is *R* = 0.66 (Spearman rank, 95%-CI: 0.40 to 0.81) indicating only a moderate correlation. This scattering of the MO values for samples with nearly the same parasitemia naturally arises, at least in part, from the fact that RMOD quantifies the amount of hemozoin in blood, which depends not only on the overall parasite density, but also on the stage distribution of the circulating parasites and the clearance rate of hemozoin from the circulation. This scattering of the MO values due to the listed factors also explains the relatively large error of the detection limit specified above.

While the MO signal shows only a moderate correlation with the parasite density, it correlates better with the estimated hemozoin content (*HZ*), which is a parameter calculated by assuming different hemozoin production rates of the different erythrocytic stages. When we use a subset of samples with clear dominance of rings and trophozoites, i.e., that contain schizonts and gametocytes in less than 3%, the hemozoin content is well approximated by *HZ* = 1 · *Ri*+ *α* · *T*, where *Ri* and *T* denote the density of rings and trophozoites, respectively, and *α* is the relative hemozoin contribution of trophozoites compared to rings. (Please note that *HZ* parameter is a weighted parasite density, which is not equal to the actual hemozoin concentration, but is linearly proportional to it.) As shown in Fig.3A., the Spearman correlation coefficient between the MO values and the corresponding *HZ* values reaches its maximum, *R* = 0.9, at *α* = 4.5 *±* 0.3. Although the assessment of the relative production rate for schizonts and gametocytes is less accurate due to their rare occurrence and the limited statistical sample size, their contribution is estimated to be approx. 12 − 20 times higher than the hemozoin content of rings. As presented in Fig.3B., for the full sample set the best Spearman correlation, *R* = 0.75 (95%-CI: 0.55 to 0.87), was achieved with the following relative hemozoin contributions of the different stages: *HZ* = 1 · *Ri* + 4.5 · *T* + 17 · (*S* + *G*), where *S* and *G* denote the density of schizonts and gametocytes in the samples, respectively.

**Figure 3.**
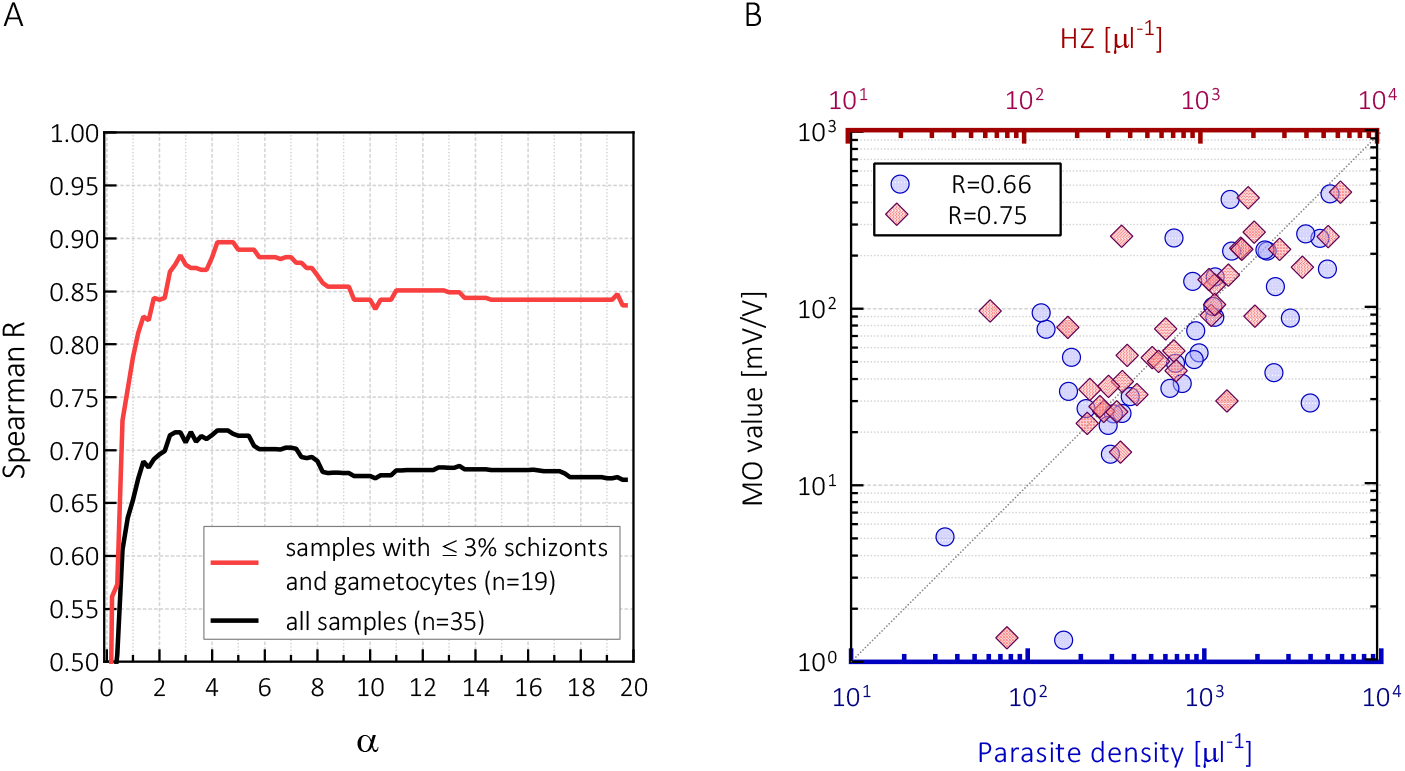
**Panel A**. The Spearman rank correlation coefficient between the MO values and the estimated hemozoin content, *HZ* = 1 · *Ri* + *α* · *T*, as the function of *α*. The red curve shows the correlation coefficient calculated for the subset of samples in which the proportion of schizonts and gametocytes was below 3% (n=19). The black curve shows the correlation coefficient for the full sample set (n=35). **Panel B**. The MO values as the function of parasite density (bottom x-axis, blue circles) and as the function of *HZ* (top x-axis, red diamonds).The latter was calculated using the value *α* = 4.5 (determined from the red curve in panel A), and also including the hemozoin contributions from schizonts and gametocytes, as described in the main text. The Spearman rank correlation coefficients for the two cases are also indicated.

## Discussion

The RMOD was able to classify all *P. vivax*-infected samples as malaria-positive with MO values typically 1 − 3 orders of magnitude higher than the cut-off level determined using samples of malaria-naïve volunteers. Furthermore, by preparing serial dilutions from the original patient samples, we found that parasite densities as low as a few parasites per a microliter of blood are still detectable by RMOD.

In our previously conducted large-scale field trial in Papua New Guinea, where expert light microscopy was used as the main reference method, the median MO value of the samples obtained from suspected patients whose light microscopy results were negative, was found to be 1.68 mV/V (IQR: 1.06 − 2.89 mV/V) [15]. This value is significantly higher than the mean MO value of the control samples in the current Thai study: 0.55 mV/V (IQR: 0.36 − 0.89 mV/V). The most plausible hypothesis to explain this observation is that a considerable portion of the patients at PNG, a high-transmission location, has residual hemozoin in the peripheral blood either due to low-density asymptomatic infections, or due to hemozoin crystals remaining in the circulation after recently treated or cleared infections. This factor increased the MO cut-off level in the PNG study and, thus, seemingly compromised the diagnostic performance.

To further elaborate on this, we make a direct comparison between the field samples from Thailand and PNG in Fig.2C. where we present the MO values for all light microscopy-positive *P. vivax* samples collected in PNG (data reproduced from [15]). This comparison reveals that the Thai and PNG data sets follow the same trend and the magnitude of the MO values in the two studies spans the same range. Most importantly, for 98% of the samples found *P. vivax*-positive in the PNG study the MO values are located above 0.94 mV*/*V, the cut-off level determined in the current study. This supports the assumption that a considerable fraction of false-positive MO detections in the PNG study are due to the high residual hemozoin level present in the population at high transmission settings. However, to firmly confirm this hypothesis further large-scale studies are crucial in low-transmission and/or elimination settings.

RMOD quantifies the concentration of hemozoin in the peripheral blood which can be present within the parasites, erythrocytes or leukocytes at the time of sampling. Since a coarse categorisation of the erythrocytic stages was carried out and the densities of the different stages were determined by expert microscopy, the hemozoin production rates of the different stages could be estimated based on the MO values of the samples. We found that trophozoites in these *in vivo P. vivax* samples contain approx. 4.5 times more hemozoin than rings, while older stages (schizonts and gametocytes) contain 12 − 20 times larger amounts than rings. These values are comparable to the relative hemozoin production rates of rings, trophozoites and schizonts reported previously for *P. falciparum* cell cultures [14, 22, 23]. From this we conclude that the actual hemozoin content in peripheral blood primarily reflects the momentary hemozoin production rate, i.e., a dynamic equilibrium is maintained between hemozoin production and clearance, as also observed in rodent infections [12, 13]. However, since RMOD could also confirm the infection in samples containing exclusively ring stages, whose hemozoin content is much lower than the hemozoin content of trophozoites and schizonts, a non-negligible hemozoin accumulation also has to occur in the peripheral blood during acute infections.

The most important requirements towards a novel method for in-field malaria diagnostics are the ease of use and high sensitivity — ideally even the detection of asymptomatic infections. However, besides the highly reliable differentiation between positive and negative cases, the quantitative assessment of disease severity might provide crucial information in certain clinical situations. In clinical practice today the parasite density is used as the primary characteristics of acute malaria infections, however, the proportion of hemozoin-containing white blood cells has also been investigated as a potential indicator of prognosis [17, 24–26]. The relation between the intraleukocytic hemozoin content and disease severity reported in former studies implies that the overall hemozoin concentration in the peripheral circulation may also serve as a useful clinical indicator [17, 24]. The quantitative nature of RMOD, realising a hemozoin-based diagnosis, can also be highly advantageous in this respect.

## Materials and Methods

### Sample collection and characterisation in Thailand

Sample collection was carried out in two field-clinics in the Kanchanaburi and Ratchaburi provinces of western Thailand from May 2014 to June 2015 (n=33), while two additional infected samples were obtained in July, 2016. Patients attending to the clinics exhibiting symptoms consistent with malaria were tested for the infection using an SD Bioline Malaria Ag P.f/Pan rapid diagnosis test kit (Standard Diagnostics Inc.) and/or via light microscopic (LM) examination of thick blood smears. Once a positive diagnosis was established, a blood sample of approximately 250 *µ*l was collected into an EDTA-containing Microtainer® (Becton Dickinson and Company) via venipuncture from consented volunteers. The blood aliquots were subsequently frozen and transported over dry ice to the Mahidol Vivax Research Unit (MVRU) of Mahidol University, Bangkok, where they were stored at −70°C until the completion of the RMOD measurements.

Approximately 40 uninfected blood samples were collected from two consented, malaria-naïve colleagues on-site at various time points over the study period for protocol optimisation and for the establishment of an uninfected baseline signal for the RMOD. These control samples were collected, frozen and stored the same way as the infected samples. Seven samples from both individuals were utilised to determine the uninfected baseline signal and the cut-off level of the study (see Fig.2A.) while the rest of the samples were used to prepare dilution series from ten infected samples (see Fig.2B.).

As a reference method for the RMOD study, a good-quality, Giemsa-stained thick blood smear was prepared from all *P. vivax*-infected samples on site, and analysed later by an expert microscopist at the central laboratory of MVRU, Bangkok. The morphological analysis of the parasites confirmed that all samples were infected exclusively with *P. vivax*. The thick blood smears were prepared using exactly one microliter of blood, thus by counting all the parasites in the total area of the smears (*×*1000 magnification) the parasite density was therefore determined directly. Furthermore, the parasites detected in the smears were assigned to stage groups (rings, trophozoites, schizonts, male gametocytes and female gametocytes) according to their morphology.

### Sample collection and characterisation in PNG

The sample collection was carried out at two health facilities in Madang, located on the north coast of Papua New Guinea where *P. falciparum* and *P. vivax* are highly endemic [27]. During the period of November, 2017–July, 2018 *n=945* blood samples were collected from suspected malaria patients and analysed by RMOD and multiple control methods, out of which *n=104* were found to be *P. vivax* monoinfections using LM as reference. Two blood slides for LM were prepared at enrolment and the LM analysis was carried out by the PNG Institute of Medical Research microscopy unit consisting of experienced, WHO-certified microscopists.

Thick blood smears were examined independently by two microscopists, for 200 thick-film fields (*×*1000 magnification) before being declared *Plasmodium*-negative. Parasite density was calculated from the number of parasites per 200–500 leukocytes (depending on parasite density) and an assumed leukocyte density of 8000 µl [28]. Slides were scored as LM-positive for an individual *Plasmodium* species if the species was detected independently by at least two microscopists.

### Ethics statements

The study conducted in Thailand received ethical clearance from the Ethics Committee at the Faculty of Tropical Medicine, Mahidol University (MUTM 2013-027-01). The study was clearly explained to all volunteers and informed consent was obtained from all participants in the study.

The study conducted in Papua New Guinea received ethical clearance by the PNG Institute of Medical Research (PNGIMR) Institutional Review Board and the PNG Medical Research Advisory Committee (MRAC, #16.45). All methods were carried out in accordance with the relevant guidelines and regulations of the named institutes.

### Magneto-optical measurements

The concept of the rotating-crystal magneto-optical setup and the underlying physical principles of hemozoin detection are described in detail in our former studies [10, 15]. Briefly, the liquid sample containing the elongated paramagnetic hemozoin crystals is inserted into the centre of a ring-shaped assembly of permanent magnets, which creates a strong uniform magnetic field at the sample position. When the magnetic ring is rotated, the crystals follow this rotation which modulates the intensity of the light passing through the sample. The modulated intensity divided by the average intensity, which has been shown to be linearly proportional to the crystal concentration, corresponds to the measured ‘MO values’ displayed in the corresponding figures in mV/V units [10].

For the RMOD analysis in Thailand, the frozen blood samples were thawed on room temperature, mixed thoroughly and 35 µl of the partially lysed blood was mixed with 315 µl of lysis solution (13 mM of NaOH and 0.03% (v/v) of Triton X-100 in distilled water) and allowed to stand for approximately 5 minutes to ensure complete lysis and homogenisation. Thereafter, 280 µl of the lysate, hereinafter referred to as optical sample, was transferred into the optical sample holders and the RMOD measurements were performed without further delay. Optical samples for RMOD measurements were prepared in duplicate per blood sample.

For the estimation of the RMOD sensitivity threshold two- or four-fold dilution series were prepared from ten infected samples. The infected samples were thawed and diluted with thawed aliquots of malaria-naïve blood samples frozen and stored together with the infected specimens. After the thorough mixing of the two components the final optical samples for the RMOD measurements were prepared as described above.

The magneto-optical measurements in Thailand and PNG were carried out with two RMOD devices that were technical replicates using the same measurement protocols. The only differences between the two RMOD experiment series were that in PNG the samples were freshly hemolysed and measured in triplicates per blood sample, while in Thailand the whole blood samples were frozen and stored until the completion of the RMOD measurements and they were measured in duplicates per blood sample.

## Data Availability

All data available upon request from the corresponding author.

## Acknowledgments

The authors would like to sincerely thank all study participants. We thank all involved health facility staff for their collaboration in these research studies. R.J.L is the recipient of an Australian National Health and Medical Research Council (NHMRC) Emerging Leadership Fellowship (GNT1173210). S.K. is a recipients of an NHMRC Career Development Fellowship (GNT1141441). A.B. and A.O. were supported by the BME-Nanotechnology and Materials Science FIKP grant of EMMI (BME FIKP-NAT) and NRDI Fund (TKP2020 IES, Grant No. BME-IE-NAT), Hungary.

## Author contributions

A.B., I. K. and A.O. developed the RMOD setup. R.J.L. and P.S. collected the patient samples in Thailand. N.M. carried out the light microscopy analysis of the Thai samples. S.K., W.N., I.M. and J.S coordinated the field studies in Thailand and PNG. A.O. carried out the RMOD measurements in Thailand.I.K., S.K. and A.O. analysed the data and wrote the manuscript. I.K. designed and supervised the project. All authors contributed to the discussion of the results.

## Dual publication statement

A portion of data originating from one of our former studies is included in the current manuscript for comparison with our new, previously unpublished results. Specifically, parasite density and magneto-optical values of *Plasmodium vivax* - infected samples collected in Papua New Guinea, which have been presented previously in Ref. [15]. The origin of the data is clearly indicated in the legend of Fig.3. and throughout the manuscript.

